# Early life seizures and epileptic spasms in *STXBP1*-related disorders

**DOI:** 10.1101/2023.06.26.23291892

**Authors:** Kim M Thalwitzer, Julie Xian, Danielle deCampo, Shridhar Parthasarathy, Jan Magielski, Katie Rose Sullivan, James Goss, Charlene Son Rigby, Michael Boland, Ben Prosser, Sarah M Ruggiero, Steffen Syrbe, Ingo Helbig

## Abstract

**Background and Objectives:** Individuals with disease-causing variants in *STXBP1* frequently have epilepsy onset in the first year of life with a variety of seizure types, including epileptic spasms. However, the impact of early-onset seizures and anti-seizure medication (ASM) on the risk of developing epileptic spasms and impact on their trajectory is poorly understood, limiting informed and anticipatory treatment, as well as trial design.

**Methods:** We retrospectively reconstructed seizure and medication histories in weekly intervals for individuals with *STXBP1*-related disorders with epilepsy onset in the first year of life and quantitatively analyzed longitudinal seizure histories and medication response.

**Results:** We included 61 individuals with early onset seizures, 29 of whom had epileptic spasms. Individuals with neonatal seizures were likely to have continued seizures after the neonatal period (25/26). The risk of developing epileptic spasms was not increased in individuals with neonatal seizures or early infantile seizures (21/41 vs. 8/16; OR 1, 95% CI 0.3-3.9, *p* = 1). We did not find any ASM associated with the development of epileptic spasms following prior seizures. Individuals with prior seizures (n = 16/21, 76%) had a higher risk to develop refractory epileptic spasms (n = 5/8, 63%, OR =1.9, 95% CI 0.2-14.6, *p* = 0.6). Individuals with refractory epileptic spasms had a later onset of epileptic spasms (n = 20, median 20 weeks) compared to individuals with non-refractory epileptic spasms (n = 8, median 13 weeks; *p* = 0.08). When assessing treatment response, we found that clonazepam (n = 3, OR 12.6, 95% CI 2.2-509.4; *p* < 0.01), clobazam (n=7, OR 3, 95% CI 1.6-6.2; *p* < 0.01), topiramate (n=9, OR 2.3, 95% CI 1.4-3.9; *p* < 0.01), and levetiracetam (n=16, OR 1.7, 95% CI 1.2-2.4; *p* < 0.01) were more likely to reduce seizure frequency and/or to maintain seizure freedom with regards to epileptic spasms than other medications.

**Discussion:** We provide a comprehensive assessment of early-onset seizures in *STXBP1*-related disorders and show that the risk of epileptic spasms is not increased following a prior history of early-life seizures, nor by certain ASM. Our study provides baseline information for targeted treatment and prognostication in early-life seizures in *STXBP1*-related disorders.

## INTRODUCTION

*STXBP1*-related disorders are one of the most common genetic diagnoses in individuals with epilepsy.^1^ *STXBP1* encodes the Syntaxin-binding protein 1 (STXBP1 or MUNC18-1), a protein of the soluble NSF attachment protein receptor (SNARE) complex which is essential for synaptic vesicle release.^2–4^ While initially identified as developmental and epileptic encephalopathy (STXBP1-DEE), *STXBP1*-related disorders are increasingly recognized as a spectrum, also including muscular hypotonia, movement disorders, tremor, and behavioral differences.^5^

Seizures are present in 90% of individuals, occur most commonly in the first year of life^5^, and are associated with a significant disease burden that impacts individuals and their caregivers^6^. Forty-two percent of individuals with *STXBP1*-related disorders have epileptic spasms, with the risk increased in individuals with protein-truncating variants (PTV) and deletions compared to missense variants.^5^ The use of certain anti-seizure medications (ASM), in particular sodium-channel blockers, is thought to provoke epileptic spasms^7^, suggesting that the choice of early ASM for early-life seizures may contribute to the development of epileptic spasms.

Currently, it is unclear whether a prior history of early-life seizures and medication use are risk factors for epileptic spasms in individuals with *STXBP1*-related disorders. Given that the genetic diagnosis can have critical implications on clinical management and is commonly given after the occurrence of early-infantile seizures, this knowledge is essential to provide optimal care with the goal to reduce the risk of epileptic spasms in individuals with *STXBP1*-related disorders.

Here, we evaluated seizure histories and medication efficacy in 61 individuals with *STXBP1*-related disorders with seizure onset during the first year of life. We aimed to identify the impact of early-onset seizures and ASMs on the risk and outcomes of individuals with epileptic spasms.

## Methods

### Study inclusion of individuals with *STXBP1*-related disorders

Individuals with *STXBP1*-related disorders were recruited through the Epilepsy Neurogenetics Initiative (ENGIN) and the Epilepsy Genetics Research Project (EGRP) at Children’s Hospital of Philadelphia (CHOP) using the following inclusion criteria: (1) a disease-causing variant in *STXBP1*, (2) seizure onset in the first year of life, and (3) sufficient data on seizure outcome. For the following analyses, individuals were excluded: (1) for analysis of trajectories of seizure types across the first year of life, individuals that were under one year of age and had no prior history of epileptic spasms, as we could not exclude that these individuals will not develop epileptic spasms eventually, and (2) for analysis of comparative effectiveness of ASMs, individuals that did not have detailed information on medication histories and changes in weekly seizure frequencies, and (3) for analysis of phenotypic correlations with variant subgroups, individuals with in-frame deletion or in-frame insertion due to unknown functional consequences.

### Phenotypic and genotypic annotation

We retrospectively reviewed clinical records to reconstruct seizure histories in weekly time intervals. Seizure types were captured using the Human Phenotype Ontology (HPO), a standardized framework for mapping clinical features.^8, 9^ The phenotype “epileptic spasms” refers to the current ILAE definition, referring to individuals with infantile epileptic spasms syndrome, consisting of epileptic spasms with or without hypsarrhythmia, mainly occurring in the first two years of life.^10^ We defined neonatal seizures as seizure onset within the first month of life. “Early infantile seizures” refers to individuals that have seizure onset after the first month of life and until the age when 90% of individuals with epileptic spasms developed epileptic spasms. Seizure frequency scores (SF) were assessed using previously published standard nomenclature derived from the Epilepsy Learning Health System and Pediatric Epilepsy Learning Health System (ELHS/PELHS standards): multiple daily seizures (>5 per day, SF score = 5), several daily seizures (2–5 per day, SF score = 4), daily seizures (SF score = 3), weekly seizures (SF score = 2), monthly seizures (SF score = 1), no seizures (SF score = 0). When assessing seizure frequency on a group level, we used the median seizure frequency scores and displayed the range of values. Individuals that (1) had a reoccurrence of epileptic spasms after initial remission, or (2) needed more than one typical ASM (ACTH, prednisolone, vigabatrin) for treatment of epileptic spasms, or (3) had a duration of epileptic spasms that lasted six weeks or longer were classified to have treatment resistant epileptic spasms.

For analysis of genotype-phenotype correlations, we included PTVs, splice site variants, frameshift variants and whole and partial gene deletions in a single group referred to as PTV/del and compared this subgroup with individuals with missense variants.

### Reconstruction of ASMs and assessment of treatment effectiveness

We manually reviewed all medical charts for the presence of ASM in the first year of life and assessed start and end dates for each medication, which was then binned in weekly intervals. If the start and wean date of medications were unclear, the first and last time a medication was used was documented. Abortive rescue medications were excluded from the analysis. We assessed the comparative efficacy of ASM by analyzing the frequency of intervals with seizure improvement coinciding with medication use compared to the frequency of intervals with seizures improvement with the use of other medications, as previously published.^5^ For example, we assessed the number of time intervals in which phenobarbital was prescribed and the change in seizure frequency during these intervals compared to the change in seizure frequency across intervals in which individuals received a medication other than phenobarbital.

### Statistical analysis

All computations were performed using the R Statistical Framework.^11–14^ We derived odds ratios using Fisher’s exact test for three assessments of comparative effectiveness and analyzed subgroups within different seizure types: (1) seizure frequency reduction (2) seizure frequency reduction or maintaining seizure freedom, defined by no seizures for consecutive weeks, and (3) maintaining seizure freedom only. Periods with unclear seizure or medication history were excluded from the analysis of ASM efficacy. Reported *P* values were two-sided, with *P* ≤ 0.05 considered statistically significant*. P* values were adjusted for multiple hypothesis testing using Bonferroni-procedure holding a false discovery rate (FDR) of 5%.

### Standard Protocol Approvals, Registrations, and Patient Consents

The study was conducted following local protocol at Children’s Hospital of Philadelphia (IRB 12226), allowing for the recruitment of individuals, acquisition of health information from electronic medical records, and publication of de-identified data. Informed consent was obtained for all individuals.

## RESULTS

### A subset of 61 individuals with STXBP1-DEE have epilepsy onset in infancy

We identified 65 individuals with *STXBP1*-related disorders and epilepsy onset in the first year of life at a major *STXBP1* specialty clinic. Of those 65 individuals, we excluded four individuals due to incomplete clinical data. Twenty-five out of 61 individuals (41%) were female. Twenty-five individuals had missense variants, ten individuals had PTVs, nine individuals had frameshift variants, eight individuals had whole or partial gene deletions, seven individuals had splice site variants, one individual had an in-frame deletion, and one individual had an in-frame insertion **(Table 1)**. For analysis of genotype-phenotype correlations, we included PTVs, splice site variants, frameshift variants and whole and partial gene deletions in a single group referred to as PTV/del (n=25) and compared this subgroup with individuals with missense variants (n=34).

**Table 1.** Cohort information.

|  |  |
| --- | --- |
| Individuals with *STXBP1*-related disorders: | 61 |
| Included in comparative effectiveness analysis: n | 52 |
| Sex |  |
| Female | 25 (41%) |
| Epilepsy types |  |
| Neonatal seizures | 26 (46%) |
| Focal-onset seizures | 48 (84%) |
| Generalized-onset seizures | 4 (7%) |
| Epileptic spasms | 29 (51%) |
| Genetic variants |  |
| Missense | 25 (41%) |
| Nonsense | 10 (16%) |
| Frameshift | 9 (15%) |
| Whole/partial gene deletions | 8 (13%) |
| Splice site | 7 (11%) |
| In-frame deletion | 1 (2%) |
| In-frame insertion | 1 (2%) |

### Epileptic spasms are present in half of individuals with seizures in the first year of life

All individuals included in our cohort had seizure onset in the first year of life. The median age at onset was 5 weeks (w) (IQR = 1-13w). The weekly proportion of individuals with seizures ranged from 17% (n=9/54, week 3) to 40% (n=23/57, week 11). For the following analyses, individuals aged younger than 12 months have been excluded, as seizure trajectories across the first year of life are unclear and seizure types as epileptic spasms might occur at the end of the first year of life. Excluded individuals had neonatal seizures (n=1), early infantile seizures (n=1), and neonatal and early infantile seizures (n=2).

Of the remaining 57 individuals, twenty-six (46%) individuals had neonatal seizures, including 18 (32%) with seizure onset in the first week of life. Focal-onset seizures were the most common seizure type (n=48, 84%) and occurred at a median age of 4.5 weeks (IQR 1-13w) with the maximum proportion at the first week of life (n=18, 32%) in addition to weeks eight through eleven of life (n=14-18/57). Four individuals (7%) had generalized-onset seizures occurring at a median age of 16 weeks (IQR = 9-29w) **(Fig. 1a)**. Twenty-nine individuals (51%) had epileptic spasms **(Table 1)**. The median age of onset was at the age of 17 weeks (IQR = 10-22w) and 90% of individuals had an onset before the age of 28 weeks. The minimum age of onset was four weeks, and the maximum age of onset was 45 weeks. Individuals were more likely to have epileptic spasms within weeks 22 and 23 (22%, n=12/55-56) and the prevalence of epileptic spasms decreased over time to 11% (6/54) by the end of the first year of life **(Fig. 1b)**. The median duration of epileptic spasms was 11 weeks (IQR = 2-22w), with a maximum duration of 39 weeks **(Fig. 1c)**.

**Figure 1.**
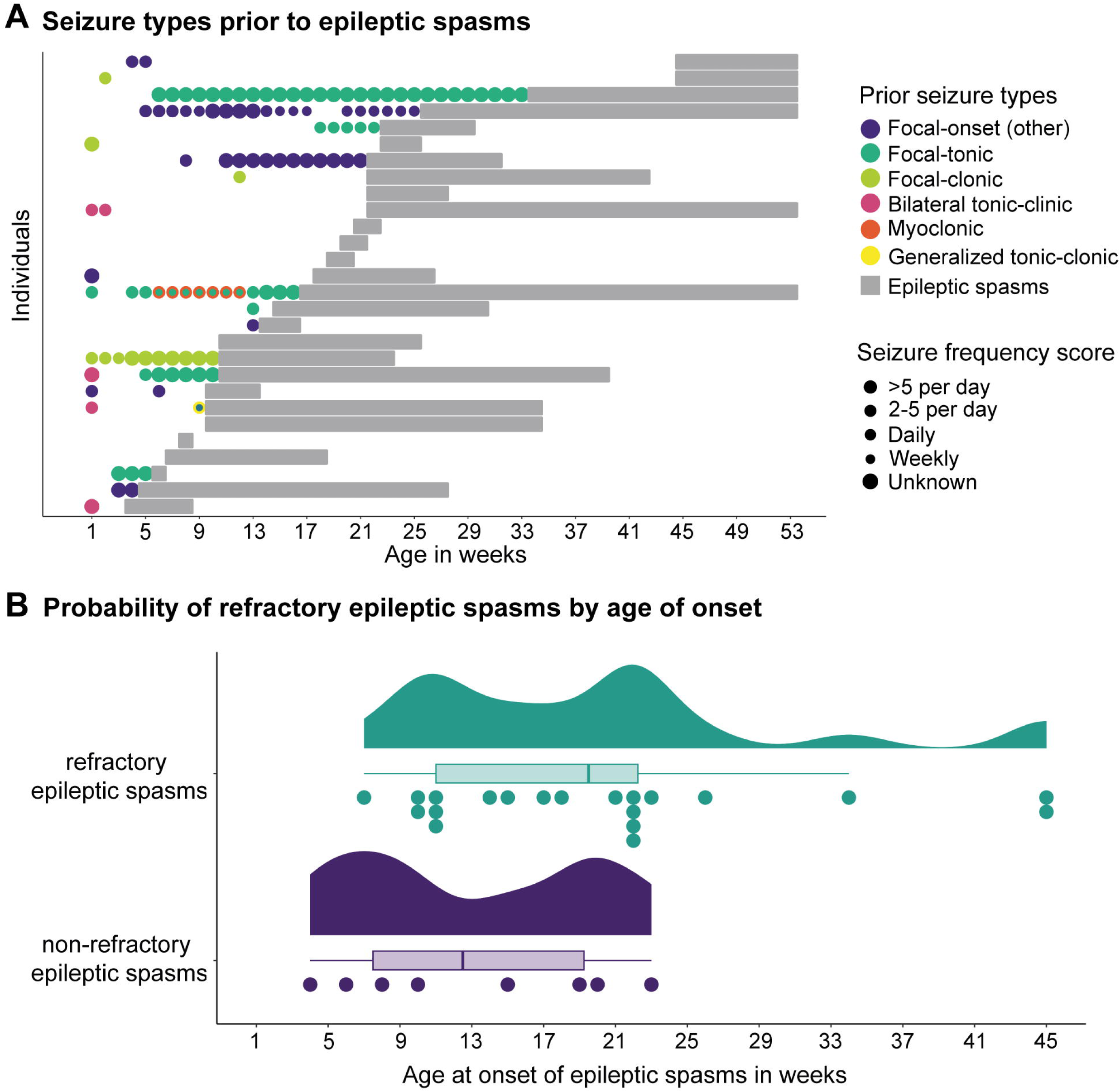
Patterns of seizures in the first year of life. **(A)** Proportion of individuals with seizures (in black) and distribution of seizure types (different colors) in weekly time intervals during the first year of life. Seizures occurred mainly in week of life one (n=18/57) and between weeks 8 and 13 (n = 20-23/57). The highlighted bars indicate the weeks with highest occurrence of focal-onset seizures (week one, eight through eleven, n=14-18/57) and epileptic spasms (weeks 22 and 23, n=12/55-56). **(B)** Distribution of seizure frequency scores across the first year of life. The grey bars display proportions of individuals with seizures for each weekly time interval. The bars with color indicate proportions of individuals with epileptic spasms at a certain frequency during each respective week. Progression in blue between weekly increments indicates decreased seizure frequency, whereas yellow indicates an increased frequency score, and grey indicates no change. **(C)** Seizure frequency scores displayed per individual for the subgroup of individuals with epileptic spasms. The grey bars indicate the presence of other seizure types in the absence of epileptic spasms.

### The risk of epileptic spasms in not increased in individuals with STXBP1-DEE and a history of early-life seizures

Next, we assessed the extent to which previous seizures impacted the risk of later developing epileptic spasms. Out of the individuals with information on seizure trajectories (n=57), 26 (46%) individuals had neonatal seizures and 34 (60%) individuals had seizures between 1 month to 7 months of age which was referred to as the “early infantile seizures” as these were seizures occurring after the neonatal period, but before the age at which 90% of individuals in our cohort developed spasms. Forty-one (72%) individuals had neonatal or early infantile seizures. Nineteen (26%) individuals had both neonatal seizures and early infantile seizures. Twenty-nine (51%) individuals had epileptic spasms.

Out of the individuals with neonatal-onset seizures, all but one individual had ongoing or recurrent seizures beyond the neonatal period **(Fig. 2a)**. Individuals with neonatal or early infantile onset of seizures did not have an increased risk of subsequently developing epileptic spasms compared to individuals without seizures before the age of seven months. Twenty-one out of 41(51%) individuals with seizures and 8/16 (50%) without seizures developed epileptic spasms (OR 1, 95% CI 0.3-3.9, *p*=1), respectively **(Fig. 2b)**.

**Figure 2.**
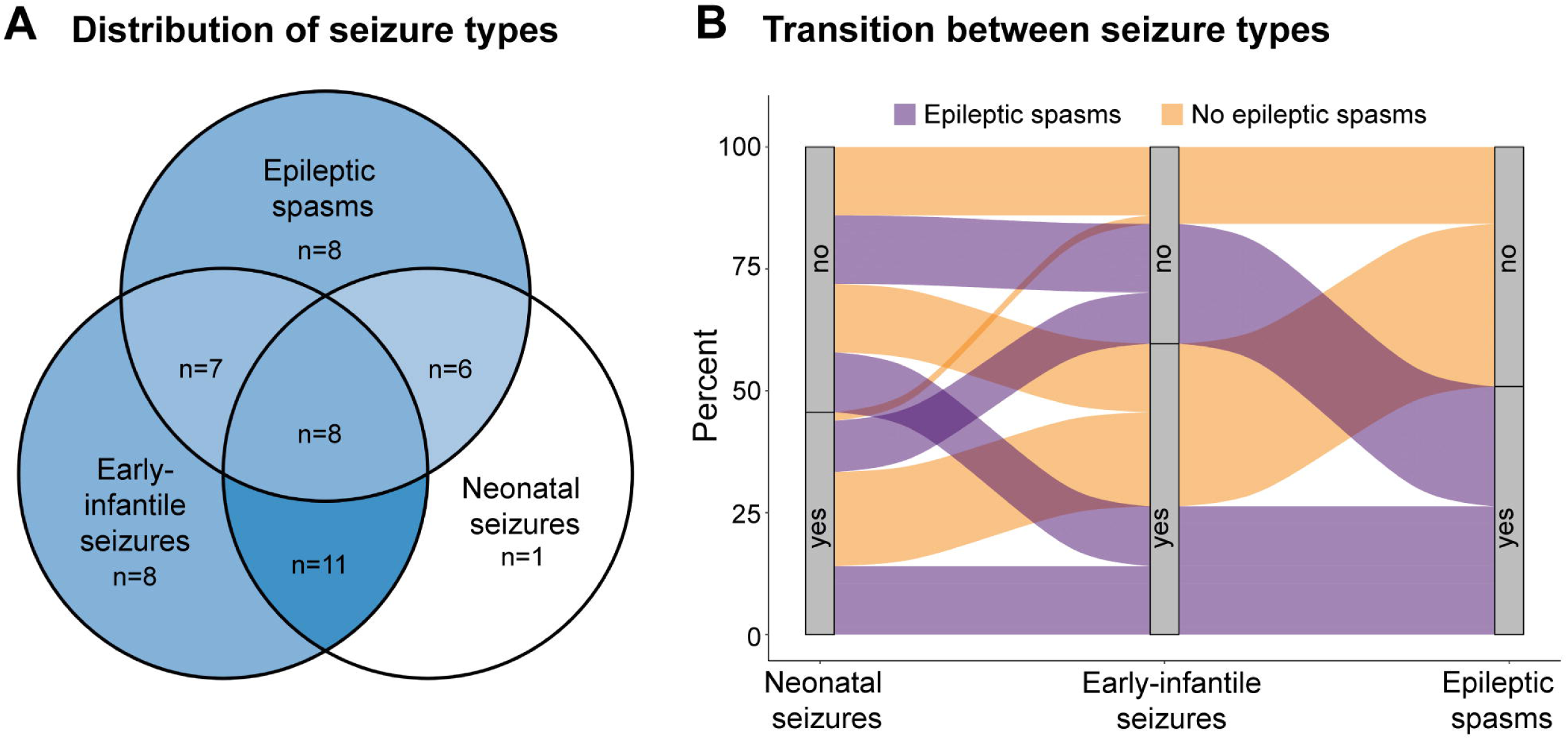
Distribution and transition between seizure types in the first year of life. **(A)** Distribution of seizure types in the first year of life. A darker color correlates with a higher number of individuals in the respective group. **(B)** Transition between seizure types in the first year of life. The grey bars show the rate of individuals with neonatal seizures, early infantile seizures (1 month – 7 months) and epileptic spasms. Progression in purple indicates individuals that will develop epileptic spasms, whereas orange indicates individuals that will not develop epileptic spasms later.

Out of 29 individuals with epileptic spasms, 21 (72%) had seizures before the onset of epileptic spasms including neonatal seizures in 14 individuals (48%), and only 8 (28%) individuals had spasms as their first presentation. Seizure types before the onset of spasms varied, including focal-onset seizures as the most frequent type (n=20/29, 69%) **(Fig. 3a)**. We did not find a correlation between seizure type and the risk for developing epileptic spasms. Furthermore, we did not find a significant difference in the distribution of PTV/del and missense variants for the seizure subgroups.

**Figure 3.**
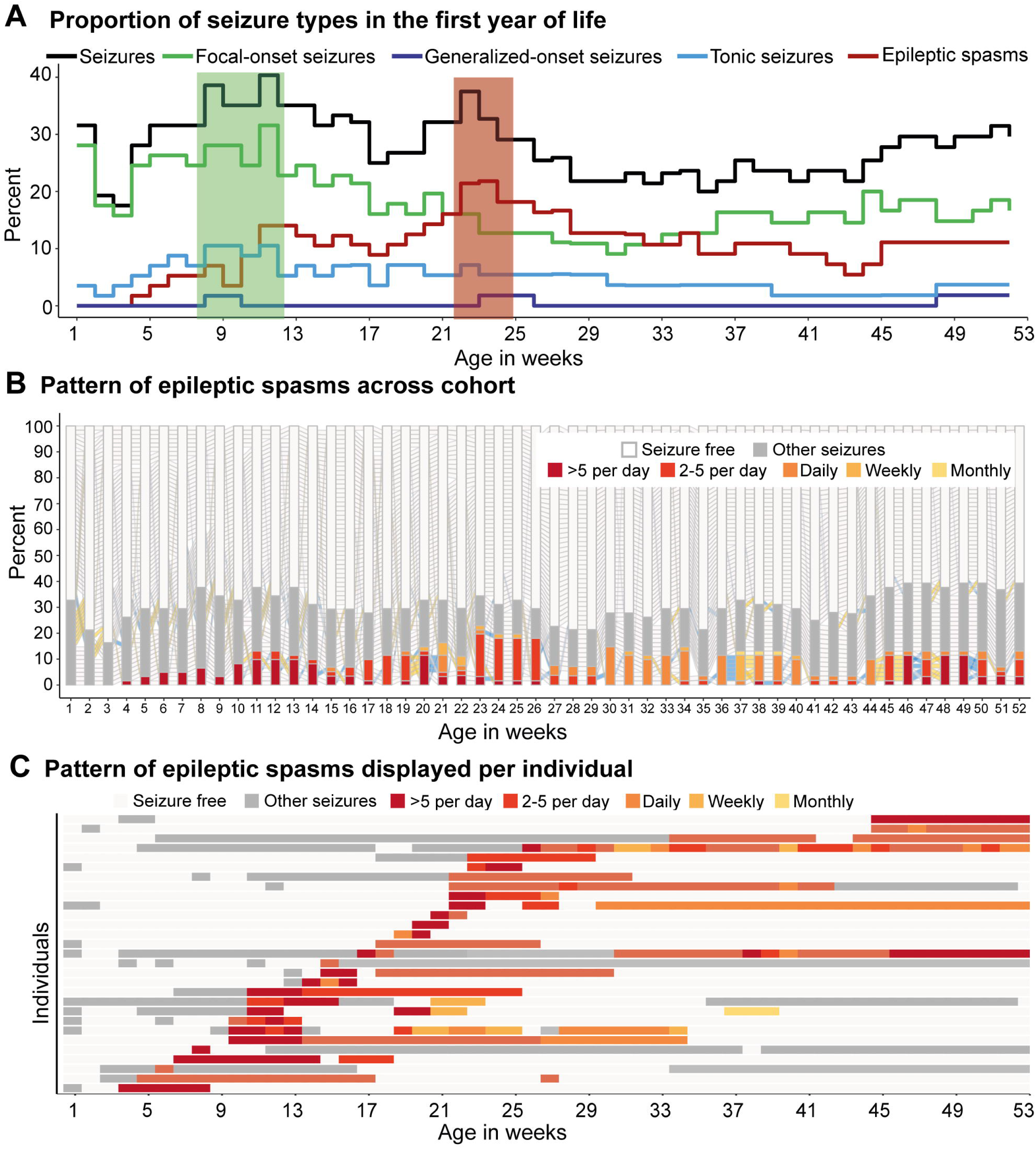
Risk factors for refractory epileptic spasms. **(A)** Seizure types before the onset of spasms are shown for each individual. In one individual, the seizure type was unknown. The larger the point size, the higher was the seizure frequency score at the time interval. For individuals with multiple seizure types, the highest seizure frequency score is displayed. **(B)** Individuals with non-refractory epileptic spasms had an earlier spasm onset (n=8, median 13w) compared to individuals with refractory epileptic spasms (n=20, median 20w, p=0.08).

### The risk for epileptic spasms in STXBP1-DEE is independent of prior anti-seizure medications

We assessed the impact of previous ASMs on epileptic spasms. First, we analyzed if single ASMs were associated with a higher risk of epileptic spasms. To this end, we stratified the number of individuals with a certain medication that developed epileptic spasms to individuals with this medication that did not develop epileptic spasms compared to individuals with other medications. We did not find an association between the use of any ASM and the risk of epileptic spasms. Furthermore, we did not find a higher risk of individuals using certain ASM within two weeks before the onset of spasms compared to individuals without epileptic spasms.

Next, we compared the risk of developing epileptic spasms with the use of sodium channel blockers including lacosamide, oxcarbazepine, phenytoin, carbamazepine, and zonisamide compared to other ASM. 3/12 (25%) individuals were treated with a sodium-channel blocker within two weeks before the onset of epileptic spasms, whereas 5 out of 32 (16%) individuals without epileptic spasms received a sodium-channel blocker (OR 1.3, 95% CI 0.18-8.3; *p*=0.7). Three individuals treated with sodium-channel blockers before the onset of epileptic spasms received oxcarbazepine, starting six, nine and 32 weeks prior to the onset of epileptic spasms. In summary, we did not find a correlation between the use of specific ASM, particularly sodium channel blockers, and the risk of epileptic spasms. None of the ASM analyzed reduced the risk of developing epileptic spasms.

### Predictors for refractory epileptic spasms

Next, we assessed risk factors for refractory epileptic spasms, including individuals that either (1) had a reoccurrence of epileptic spasms after initial remission or (2) needed more than one typical ASM for treatment of epileptic spasms (defined as ACTH, prednisolone, vigabatrin) for treatment of epileptic spasms or (3) had a duration of epileptic spasms that lasted six weeks or longer. Out of 29 individuals with epileptic spasms, 15 (52%) required more than one typical ASM, 13 (45%) had a duration of epileptic spasms of six weeks or longer and nine had a reappearance of epileptic spasms after an initial remission. In total, 21 out of 29 (72%) individuals had refractory epileptic spasms.

When stratifying by variant subgroups, we did not find an association of variant type with the risk for refractory spasms, likely due to small numbers. Nominally, individuals with missense variants had an increased risk for refractory epileptic spasms (n=8/11, 73%) compared to individuals with PTVs/del (n=12/17, 71%; OR 1.1, 95% CI 0.16-9.2, *p*=1), but this finding was not significant. Individuals with resolving epileptic spasms had an earlier spasm onset (n=8, median 13w) compared to individuals with refractory epileptic spasms (n=20, median 20w, *p*=0.08) **(Fig. 3b)**.

Finally, we compared the impact of early-life seizures on the risk of refractory epileptic spasms. Individuals with prior early-life seizures were more likely to have refractory epileptic spasms (n=16/21, 76%) than individuals with epileptic spasms as the first seizure type (n=5/8, 63%, OR =1.9, 95% CI 0.2-14.6, *p*=0.6). In summary, a later age at onset of epileptic spasms and previous early-life seizures were associated with a risk for refractory spasms.

### The effectiveness of ASM and ketogenic diet differs between neonatal and early-infantile seizures

We analyzed the relative effectiveness of ASMs across different seizure types and ages (0-3 months, 3-6 months, 6-9 months, 9-12 months, and across all ages). For this analysis, we excluded nine individuals, as detailed information on seizure frequencies and medication histories over time was not available. Individuals with focal-onset seizures in the first three months of life were primarily treated with phenobarbital (n=22/25, 88%) and levetiracetam (n=17/25, 68%). We did not find superiority of any medication in reducing seizure frequency of focal-onset seizures or maintaining seizure freedom across this age span.

Across the first year of life, levetiracetam was more likely to reduce frequency of focal-onset seizures than all other medications (n=25, OR 2.8, 95% CI 1.5-5.2; *p* < 0.01). The ketogenic diet (n=3, OR Inf, 95% CI 1.3-Inf; *p*=0.02), vigabatrin (n=10, OR 15.4, 95% CI 4.2-129.4; *p* < 0.01) and levetiracetam (n=30, OR 2.5, 95% CI 1.9-3.4; *p*<0.01) were more likely to maintain seizure freedom compared to all other medications **(Supplementary Fig. 1)**. Results of comparative effectiveness analyses across all seizure types and age spans are shown in **Supplementary Fig. 2**.

### The analysis of medication effectiveness for epileptic spasms favors ACTH, clobazam, levetiracetam, topiramate, and prednisolone

We were able to reconstruct detailed seizure and medication histories for 25 individuals with epileptic spasms. Various ASMs as well as the ketogenic diet were used to treat epileptic spasms in the first year of life, including most often ACTH (n=18), levetiracetam (n=17), prednisolone (n=13), vigabatrin (n=12), phenobarbital (n=11), topiramate (n=10), and clobazam (n=7) **(Fig. 4a)**. The median number of distinct ASMs including ketogenic diet was four, with a maximum of eight different ASMs per individual. 22 individuals received three or more different ASMs during the first year of life, indicating a drug-resistant epilepsy. Individuals were treated with up to seven medications at once **(Fig.4b)**. We found that ACTH (n=12, OR 1.9, 95% CI 0.8 - 4, *p*=0.1), clobazam (n =5, OR 1.3, 95% CI 0.4 – 3.9, *p*=0.6), levetiracetam (n=13, OR 1.2, 95% CI 0.7 - 2.2, *p*=0.5), topiramate (n=8, OR 1.1, 95% CI 0.5 –2.6, *p*=0.8), and prednisolone (n=10, OR 1.1, 95% CI 0.5 – 2.4, *p*=0.9), were more effective in reducing seizure frequency than compared to all other medications **(Fig. 4c)**.

**Figure 4.**
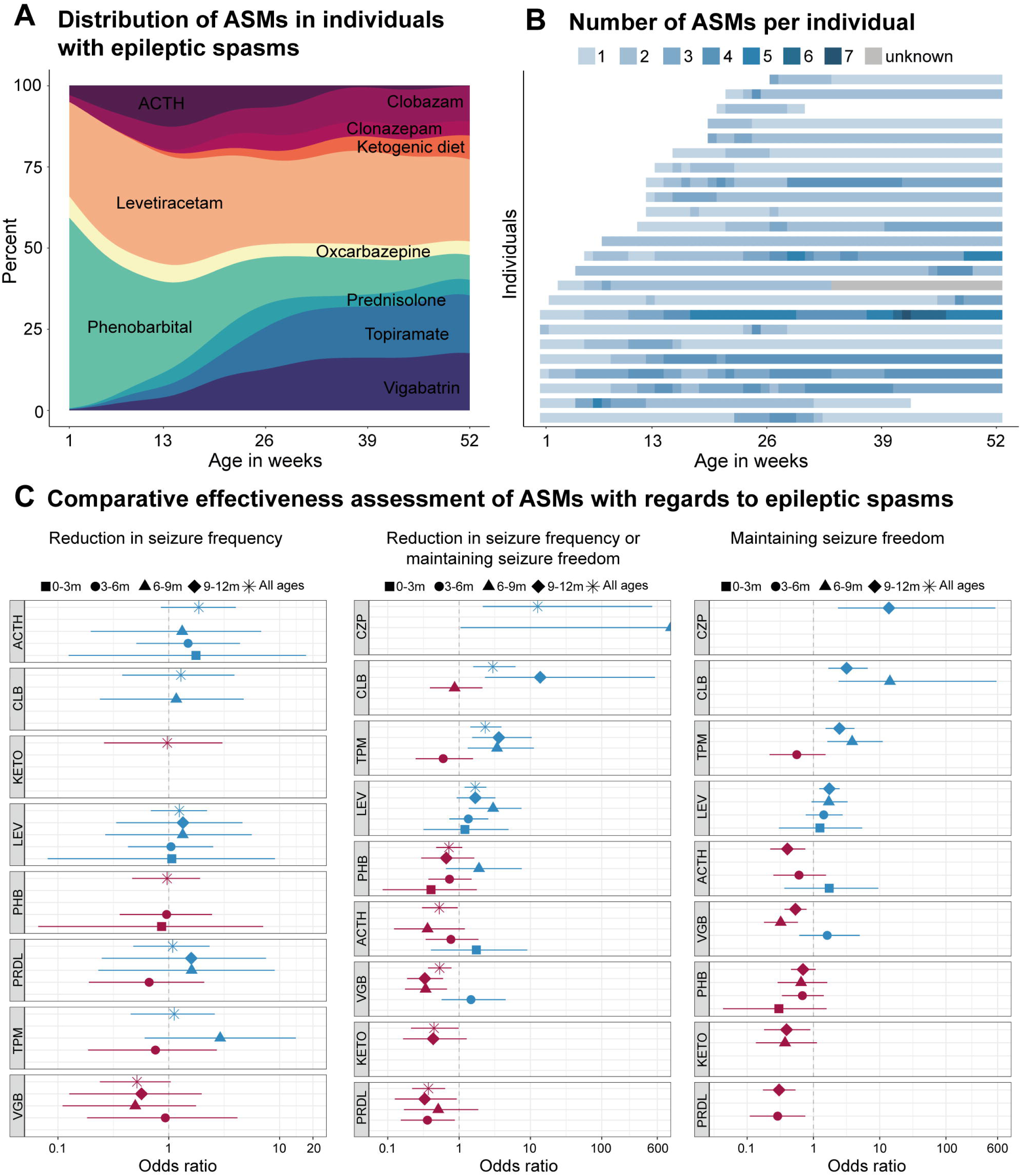
Comparative effectiveness assessment of ASM with regards to epileptic spasms. Treatment response of 25 individuals with epileptic spasms. The most frequently used ASMs were ACTH (n=18), levetiracetam (n=17), prednisolone (n=13), vigabatrin (n=12), phenobarbital (n=11), topiramate (n=10), and clobazam (n=7). **(A)** Relative distribution of ASMs and ketogenic diet in the first year of life. **(B)** Absolute number of ASMs or ketogenic diet displayed per individual across the first year of life. **(C)** Relative efficacy of ASMs and ketogenic diet in reducing frequency of epileptic spasms. **(D)** Relative efficacy of ASM and ketogenic diet in reducing seizure frequency or maintaining seizure freedom. **(E)** Relative efficacy of ASM and ketogenic diet in maintaining seizure freedom. Displayed are treatments used in three or more individuals.

We found that clonazepam (n=3, OR 12.6, 95% CI 2.2-509.4; *p*<0.01), clobazam (n=7, OR 3, 95% CI 1.6-6.2; *p*<0.01), topiramate (n=9, OR 2.3, 95% CI 1.4-3.9; *p*<0.01), and levetiracetam (n=16, OR 1.7, 95% CI 1.2-2.4; *p*<0.01) were more likely to decrease the seizure frequency or to maintain seizure freedom than other medications **(Fig. 4c)**. These medications were also significantly more likely to maintain seizure freedom compared to all other medications **(Fig. 4c)**.

## DISCUSSION

*STXBP1*-related disorders are among the most common monogenic causes for early-onset epilepsy, with epileptic spasms reported in 42% of individuals.^5^ In broader cohorts of individuals with epilepsy, epileptic spasms are associated with a high disease severity and a poor prognosis.^15^ In this study, we aimed to identify risk factors that were associated with the onset and outcome of epileptic spasms across individuals with causative variants in *STXBP1*. As a genetic diagnosis is increasingly made rapidly after the onset of seizures, knowledge regarding transition and risk of seizure types is essential to improve targeted clinical care of individuals with *STXBP1*-related disorders and to provide a prognosis of the overall disease trajectory.

Our study has several main findings: first, we found that epileptic spasms account for a major proportion of seizures in the first year of life, occurring with a wide range for age of onset and duration of spasms. The risk of developing epileptic spasms is independent of prior seizures and medications. Individuals with neonatal or early infantile epilepsy were not at higher risk to develop epileptic spasms compared to individuals with no epilepsy in the first seven months of life: we found that half of the individuals with neonatal seizures later had epileptic spasms. However, only one individual with neonatal seizures had no additional seizures in the first year of life, indicating a higher general risk for further seizures in individuals with neonatal seizures and the need for close observation of this subgroup in the clinical setting. Most individuals with epileptic spasms either had neonatal seizures or/and early infantile seizures before the onset of spasms (72%, n=21/29). The high prevalence of prior seizures in individuals with epileptic spasms aligns with findings from other genetic etiologies. For example, 88% of individuals with epileptic spasms due to *CDKL5* deficiency were reported to have seizures prior to the onset of epileptic spasms.^16^

Second, we did not find associations between any ASM and an increase or decrease in the risk for epileptic spasms. Compared to previous studies analyzing epileptic spasms in a broader cohort, we did not find that sodium-channel blockers provoke epileptic spasms.^7^ However, we did also not identify any ASM associated with a lower risk of epileptic spasms. This finding may be explained by the lack of specificity of ASM on the function of STXBP1. This finding is in contrast to epileptic encephalopathies due to other etiologies, for example, contraindication of sodium channel blockers in individuals with loss-of function variants in genes encoding sodium channels such as *SCN2A* or *SCN1A*. However, future prospective studies are needed to further evaluate medication use in *STXBP1*-related disorders.

Third, we found that epileptic spasms in *STXBP1*-related disorders are refractory in 72% of individuals (n = 21/29). This rate is higher compared to large cohorts of individuals with epileptic spasms due to various or unknown etiologies.^17, 18^ However, in other genetic etiologies, individuals with epileptic spasms also responded poorly to standard treatment of epileptic spasms: individuals with *CDKL5* deficiency have been reported to have refractory epileptic spasms in 96% compared to 53% of individuals of the National Infantile Spasms Consortium.^16^ We found that individuals with prior seizures in the first few months of life as well as individuals with a later onset of epileptic spasms were more likely to develop refractory epileptic spasms. The duration of spasms was increased in individuals with previous seizures, indicating a higher seizure burden and more overall severe disease presentation compared to individuals with spasms as the first seizure type. Interestingly, we did not identify any correlation between development of refractory spasms epileptic spasms or refractory spasms by variant type. However, our previous work indicated that the general risk for epileptic spasms is increased in individuals with protein truncating variants and deletions.^5^ This information provides granular insight into factors predictive of treatment-resistant spasms, which stands to be critical for individualized care and tailored treatment strategies. Following this subgroup of individuals will be critical in future natural history studies, particularly with respect to treatment strategies.

Our study has several limitations due to its design. First, we were not able to include all individuals in our study across all analyses due to availability of clinical and medication data. However, as we manually reviewed all information from medical charts, we aimed to improve the overall data quality by excluding individuals with incomplete data. Second, our retrospective study reports descriptive associations based on real world data, and no causal relations can be inferred. Since information from retrospective modeling is essential to design future prospective studies, our study provides important foundational information for design and endpoint measurements.

In conclusion, we show that the risk of epileptic spasms is independent of previous history of early-life seizures and medication use. However, prior seizures and later onset increases the first for refractory spasms. We provide clinical insight on the interplay between seizures and medication response in the first year of life in *STXBP1*, which has critical implications in the care of individuals with *STXBP1*-related disorders and providing a prognosis for families.

## Data Availability

All data produced in the present study are available upon reasonable request to the authors.

## Acknowledgements

We would like to thank the individuals and families with STXBP1-related disorders who participated in our study.

## Funding

I. Helbig was supported by The Hartwell Foundation through an Individual Biomedical Research Award. This work was also supported by the National Institute for Neurological Disorders and Stroke (R01 NS127830-01A1, R01 NS131512-01, and K02 NS112600), the Eunice Kennedy Shriver National Institute of Child Health and Human Development through the Intellectual and Developmental Disabilities Research Center (IDDRC) at Children’s Hospital of Philadelphia and the University of Pennsylvania (U54 HD086984), and by intramural funds of the Children’s Hospital of Philadelphia through the Epilepsy NeuroGenetics Initiative (ENGIN). Research reported in this publication was also supported by the National Center for Advancing Translational Sciences of the National Institutes of Health under Award Number UL1TR001878. This project was also supported in part by the Institute for Translational Medicine and Therapeutics’ (ITMAT) Transdisciplinary Program in Translational Medicine and Therapeutics at the Perelman School of Medicine of the University of Pennsylvania. The study also received support through the EuroEPINOMICS-Rare Epilepsy Syndrome (RES) Consortium, which provided the capacity for exome sequencing, by the German Research Foundation (HE5415/3-1 to I.Helbig) within the EuroEPINOMICS framework of the European Science Foundation, by the German Research Foundation (DFG; HE5415/5-1, HE5415/6-1 to I.H.) by the DFG/FNR INTER Research Unit FOR2715 (We4896/4-570 1, and He5415/7-1 to I.H.), and by the Genomics Research and Innovation Network (GRIN, grinnetwork.org). S.M.R was supported by an individual grant from the STXBP1 disorders foundation. S.Syrbe was funded by Dietmar-Hopp-Stiftung.

## Competing interests

All other authors do not declare any competing interests.

## Abbreviations

ASM: anti-seizure medication
CLB: clobazam
CZP: clonazepam
KETO: Ketogenic diet
LEV: levetiracetam
OXC: oxcarbazepine
PHB: phenobarbital
PRDL: prednisolone
SF-score: seizure frequency score
TPM: topiramate
VGB: vigabatrin

## Notes

### Competing Interest Statement

The authors have declared no competing interest.

### Author Declarations

IRB of the Children's Hospital of Philadelphia gave ethical approval for this work (IRB 12226).

