## Supplemental Figures for "Early life seizures and epileptic spasms in *STXBP1*-related disorders"

### Supplementary Material

**Supplementary Figure 1:** Comparative effectiveness assessment of ASM with regards to focal onset seizures.

**Supplementary Figure 2:** Comparative effectiveness assessment of ASM with regards to seizures.

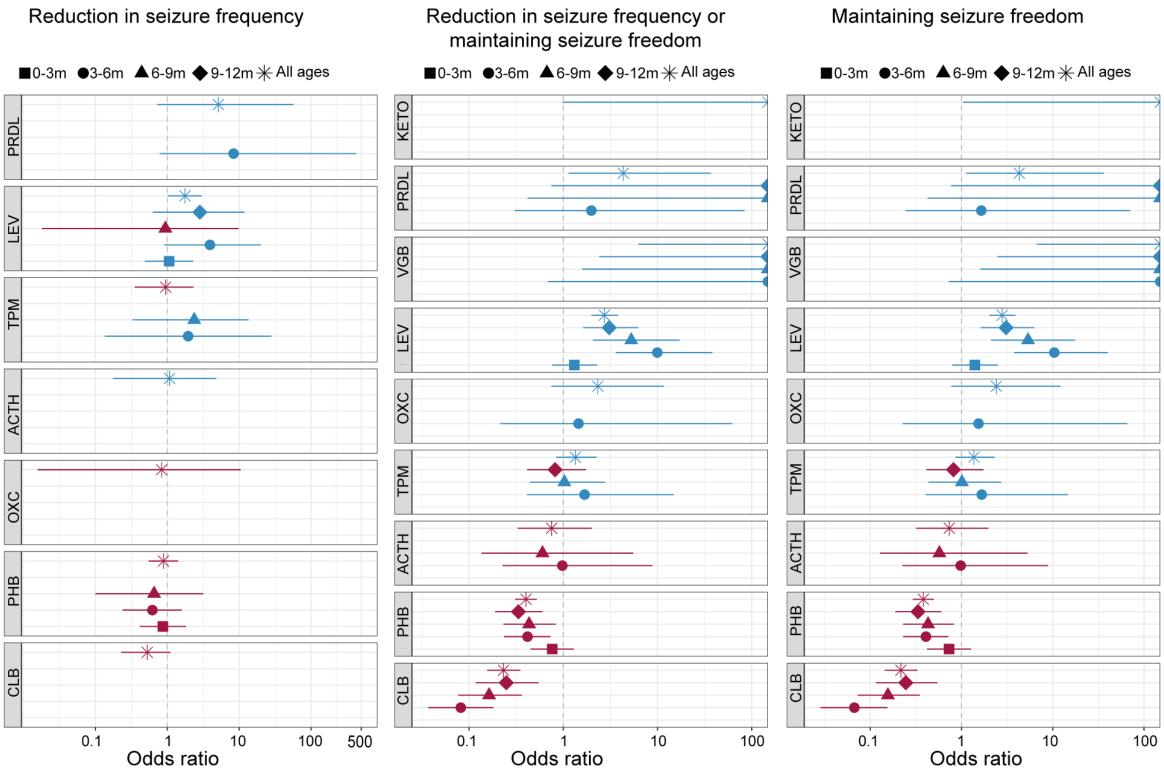


**Supplementary figure 1. Comparative effectiveness assessment of ASM with regards to focal onset seizures.** Analyzed were focal onset seizures across 44 individuals across all ages. The most often used ASMs were levetiracetam (n=32), phenobarbital (n=29), ACTH (n=14), topiramate (n=12), prednisolone (n=11), and vigabatrin (n=11).

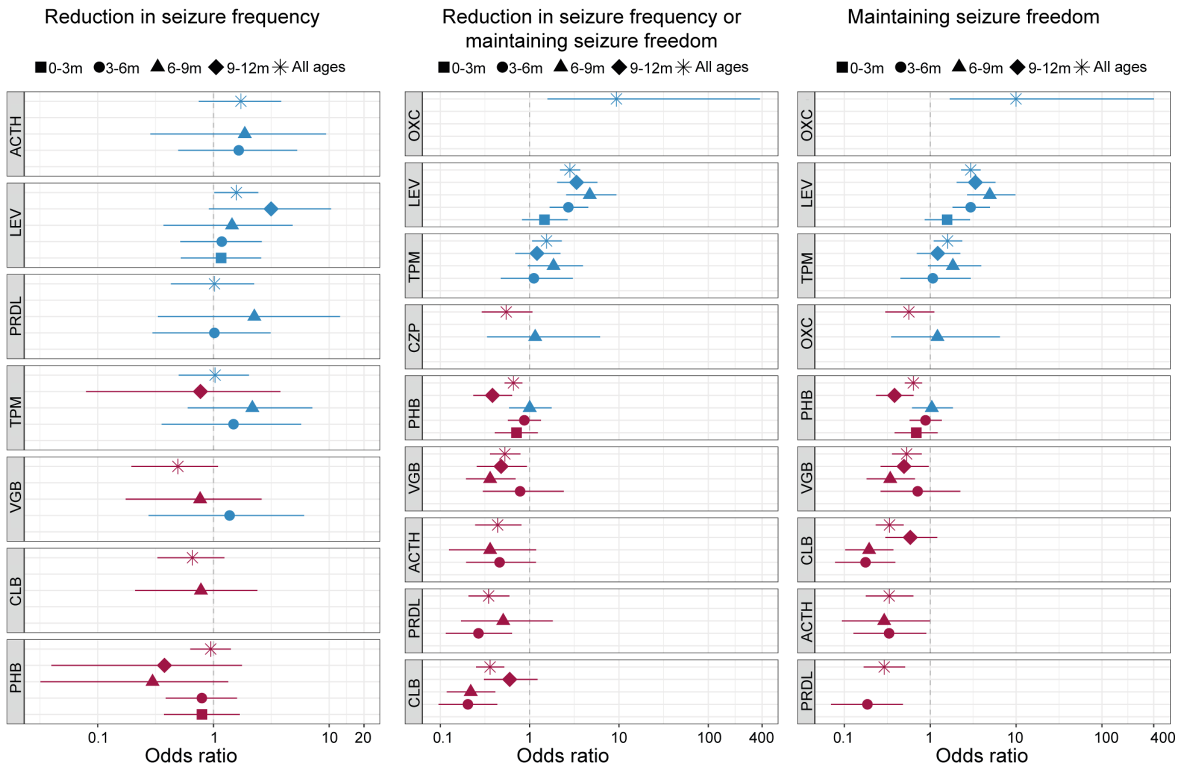


**Supplementary figure 2. Comparative effectiveness assessment of ASM with regards to seizures.** Analyzed were seizures, independent of type, across 52 individuals across all ages. The most often used ASMs were levetiracetam (n=35), phenobarbital (n=30), ACTH (n=18), topiramate (n=14), prednisolone (n=13), and vigabatrin (n=12).
